# MERS-Coronavirus across Kenya: a spatial examination of social and environmental drivers

**DOI:** 10.1101/2023.11.14.23298516

**Authors:** Ted J. Lawrence, Geoffrey Kangogo, Avery Fredman, Sharon L. Deem, Eric M. Fèvre, Ilona Gluecks, James D. Brien, Enbal Shacham

## Abstract

Climate and agricultural land-use change have increased the likelihood of infectious disease emergence and transmissions, but these drivers are often examined separately as synergistic effects are ignored. Further, seldom are the influence of climate and agricultural land use on emerging infectious diseases examined in a spatially explicit way at regional scales.

Our objective in this study was to spatially examine the climate, agriculture, and socio-demographic factors related to agro-pastoralism that can influence the prevalence of Middle East Respiratory Syndrome coronavirus (MERS-CoV) in dromedary camels across northern Kenya. Our research questions were: 1) how has MERS-CoV in dromedary camels varied across geographic regions of northern Kenya, and 2) what climate, agriculture, and socio-demographic factors of agro-pastoralism were spatially related to the geographic variation in MERS-CoV cases? To answer our questions, we analyzed the spatial distribution of historical cases of serological evidence of MERS-CoV at the county level and applied spatial statistical analysis to examine the spatial relationships of the MERS-CoV cases between 2016 and 2018 to climate, agriculture, and socio-demographic factors of agro-pastoralism.

Regional differences in MERS-CoV cases were spatially correlated with both social and environmental factors and highlight the complexity in the distribution of MERS-CoV in dromedary camels across Kenya.

## 1. Introduction

A combination of interacting social and environmental factors often drives the emergence and spread of infectious diseases (Heffernan, 2018). In particular, climate and agricultural land use intersect in ways that influence environmental conditions (Brodie, 2016), which can trigger the emergence and spread of infectious diseases. Specifically, ambient temperature affects infection rates, reproduction, and incubation time of pathogens, with higher temperatures accelerating pathogen maturation (Baker et al., 2022)(Semenza et al., 2022). Further, agriculture for food production has been associated with more than 25% of all infectious diseases and more than 50% of all zoonotic infectious diseases that have emerged in humans (Rohr et al., 2019). Lastly, the loss of biodiversity and the ensuing loss of host heterogeneity due in part to climate and agricultural land-use change has been linked to disease susceptibility and transfer (Heffernan, 2018).

In East Africa, changes in climate (i.e., hotter and drier trends) and agricultural land-use change have increased the likelihood of infectious disease emergence and transmission, such as Ebola virus, Flaviviruses, Usutu viruses, Chikungunya and O’nyong-nyong viruses, Bunyaviruses, and Rift Valley Fever and Crimean-Congo haemorrhagic viruses (Duygu et al., 2018; Fanelli & Buonavoglia, 2021; Fenollar & Mediannikov, 2018; Muturi et al., 2023; Pandit et al., 2022). Further, a combination of climate, agricultural and economic changes are supporting the spread of emerging pathogens from East Africa into the Middle East and Europe (Ryan et al., 2019; Xiao et al., 2015). Still, East Africa is considered one of the most at-risk regions in Africa to the impacts of climate change as the livelihoods of a large proportion of the region’s population depends on rain-fed agriculture (Serdeczny et al., 2017). Agro-pastoralists who depend on both livestock keeping and rain-fed crop production are considered the most vulnerable groups to climate change (Hughes & Anderson, 2020). Concurrently, population growth rates in East Africa are among the highest in the world, which increases the pressure for land conversion, and specifically for the expansion of cropland that encroaches on wildlife habitat (Bullock et al., 2021). In all, hotter and drier conditions, population growth, cropland expansion, and encroachment on wildlife habitats exacerbates infectious disease emergence and transmission (Lee-Cruz et al., 2021).

Despite the synergistic influence that climate and agricultural land use has on emerging infectious diseases, these drivers are often empirically examined separately and the potential synergistic effects are often missed (Brodie, 2016). Seldom, the influence of climate and agricultural land use have been examined related to emerging infectious diseases in a spatially explicit way at regional scales. This is especially true for Middle East Respiratory Syndrome coronavirus (MERS-CoV) that is prevalent in dromedary camels across Kenya where climate and agricultural land-use change are conspicuous (Lawrence et al., 2023a,b).

MERS-CoV is an infectious zoonotic disease that in humans targets the lower respiratory tract and can lead to multi-organ failure, resulting in death. Dromedary camels have been shown to be a natural reservoir of the virus from where spill-over to humans can occur (Adney et al, 2014). The infection is spread when a person comes into close contact with an infected dromedary camel, or possibly when a person consumes contaminated camel products such as milk and meat. Humans can also spread the virus to each other through very-close contact with infected individuals, similar to the current SARS-CoV-2 transmission (Aguanno et al., 2018).

The disease was first detected in humans in 2012 in Saudi Arabia (WHO, 2019). MERS-CoV has spread globally with more than 2,000 human infections resulting in nearly 850 identified deaths in 27 countries across North America, Europe, Asia, and Africa as of December 2019; while no new morbidity and mortality have been reported in the past several years (WHO, 2018). While no human infections have been documented in Kenya previously, the biophysical environment provides an opportunity to predict the conditions in which this may occur. Infection rates among and between camels and humans have been investigated throughout the Middle East (Reeves, 2015), Africa (Gikonyo et al., 2018; Miguel et al., 2017), and Asia (Saqib et al., 2017), which has provided preliminary mapping of infection risk. Since the discovery of MERS-CoV, serological and molecular evidence have demonstrated that the virus in dromedary camels is genetically similar to the one occurring in humans confirming the hypothesis that dromedary camels are the primary transmission reservoirs, which sheds the virus in high numbers and likely serve as reservoirs for human infections (Adney et al, 2014).

Our objective in this study was to spatially examine the climate, agriculture, and socio-demographic factors related to agro-pastoralism that can influence the prevalence of MERS-CoV in dromedary camels across northern Kenya. Our research questions were: 1) how has MERS-CoV in dromedary camels varied across geographic regions of northern Kenya, and 2) what climate, agriculture, and socio-demographic factors of agro-pastoralism were spatially related to the geographic variation in MERS-CoV cases? To answer our questions, we analyzed the spatial distribution of historical cases of serological evidence of MERS-CoV at the county level and applied spatial statistical analysis to examine the spatial relationships of the MERS-CoV cases between 2016 and 2018 to climate, agriculture, and socio-demographic factors of agro-pastoralism.

## 2. Study Site, Data Description, and Methods

### 2.1 Study site: Kenya

Located in East Africa, Kenya comprises 8 regions and 47 counties that were established through the revised constitution of Kenya in 2010 (Appendix A). Kenya’s lands are categorized as predominantly arid or semi-arid (located in the Northern Rift Valley, Eastern, Northeastern and Coastal regions) with only 15% suitable for agricultural production and roughly 80% being rangelands for the population of roughly 50 million humans (Koeva et al., 2020). Despite the relative aridity, Kenya’s arid and semi-arid regions support about 25% of Kenya’s human population, 60% of the livestock population that mostly involves pastoralism, and the largest proportion of wildlife (Ngugi & Nyariki, 2005). Further, Kenya is home to Africa’s third largest population of dromedary camels, which play a vital role in food security (Hughes & Anderson, 2020). Smallholder farmers dominate the livestock sector in Kenya with three main livestock production systems: pastoral; dairying; and ranching (Cecchi et al., 2010). Additionally, agricultural and livelihood practices in Kenya are tightly linked to agro-climatic zones (ACZs), which are the delineation of landscapes into regions with relatively homogeneous and contiguous areas based on similar climate characteristics (Boitt et al. 2014; Kogo et al. 2021; Lawrence et al., 2023a,b; (Recha, 2019)). Primarily, the ACZs in Kenya represent a temperature gradient from alpine, to temperate, to tropical regions, and a moisture gradient from humid to arid regions (Gikonyo et al., 2018).

### 2.2 Data description

The number of MERS-CoV cases in dromedary camels across northern Kenya were abstracted from six previous studies published between 2014 and 2020 (Appendix B). Each of the six studies tested for antibodies to MERS-CoV in dromedary camels and reported the results at the county level (Table 1). All of the cases of MERS-CoV in dromedary camels across northern Kenya that were used in this study were from between 1992-2018, those cases from 1992 included archived serosamples. Socio-demographic characteristics and agriculture data were from the Kenya Population and Housing Census, the Socio-Economic Atlas of Kenya, 2^nd^ edition (Wiesmann et al., 2016), and Kenyan Statistical Abstracts (KNBS, 2016, 2017, 2018).

**Table 1.** Number of samples and seropositivity of MERS-CoV cases by region, county, and year in Kenya

| Year(s) | Region | County | Samples | Seropositivity |
| --- | --- | --- | --- | --- |
| 1992 | Rift Valley | Laikipia | 22 | 5% |
| 1996 | Rift Valley | Laikipia | 37 | 5% |
| 1998 | Eastern | Isiolo | 12 | 17% |
| 1998 | Rift Valley | Laikipia | 50 | 0% |
| 1999 | Eastern | Marsabit | 41 | 78% |
| 1999 | Rift Valley | Laikipia | 175 | 18% |
| 1999 | Rift Valley | Turkana | 50 | 14% |
| 2000 | Eastern | Variable | 73 | 53% |
| 2000 | Rift Valley | Laikipia | 56 | 4% |
| 2007 | Rift Valley | Baringo | 28 | 0% |
| 2008 | Northeastern | Mandera/Wajir | 162 | 56% |
| 2008 | Eastern | Marsabit | 21 | 57% |
| 2013 | Eastern | Marsabit | 7 | 100% |
| 2013 | Rift Valley | Laikipia | 375 | 42% |
| 2016-2017 | Eastern | Isiolo | 403 | 78% |
| 2016-2017 | Eastern | Marsabit | 370 | 74% |
| 2016-2017 | Rift Valley | Turkana | 417 | 68% |
| 2016-2017 | Rift Valley | Laikipia | 181 | 15% |
| 2016-2018 | Eastern | Marsabit, Isiolo, Samburu | 293 | 80% |
| 2016-2018 | Rift Valley | Turkana, Baringo, W. Pokot | 156 | 49% |
| 2018 | Eastern | Marsabit | 493 | 76% |

Climate data were from Weather and Climate - The Global Historical Weather and Climate Data for Kenya (WC-Kenya 2023). The socio-demographic, agriculture, and climate-related data were contemporaneous with the MERS-CoV cases examined between 2016 and 2018. The socio-demographic, agriculture, and climate-related variables tested as independent variables are shown in Appendix C. The georeferenced county boundaries of Kenya were from openAFRICA (2015). The georeferenced county boundaries of Kenya were from CARTO (2016).

### 2.3 Data analysis, model evaluation and selection

We analyzed the data via a combination of non-spatial and spatial analyses using R (R Core Team, 2013), as summarized in Figure 1. Initially, we mapped MERS-CoV cases at the county level in northern Kenya using ArcGIS Pro 2.9.2 (ESRI 2022) and examined seropositivity relative to the total samples across the region, upper Rift Valley, Eastern, and Northeastern regions during the study period. In the upper Rift Valley region, we included the counties of Turkana, Laikipia, Baringo, and W. Pokot. In the upper Eastern region, we included the counties of Marsabit, Isiolo, and Samburu. In the upper Northeastern region, we included the counties of Mandera and Wajir. We then formally investigated and quantified spatial correlation of and between variables using variographic analysis, which decomposes the spatial variability of observed variables among distance classes. In this process, first, we examined the spatial autocorrelation of MERS-CoV cases. Next, we examined spatial correlation between MERS-CoV cases and a) socio-demographic variables related to agro-pastoralism, b) agriculture, and c) climate variables (Appendix C). Given that much of the socio-demographic data were from after 2010 and that the MERS-CoV data in the upper Northeastern region were only from 2008, we focused the spatial correlation analysis of MERS-CoV on 2016 through 2018 in the upper Rift Valley and upper Eastern Regions. Further, we distributed the data evenly between the northern and southern parts of each county to satisfy the practical rule that a variogram should only be applied over a specified distance for which the number of pairs is greater than 30, and because the data were not further spatially identified (Journel and Huijbregts, 1978). We fitted the data using spherical and exponential models and used a distance of between 250 to 350 km, which was deemed conservative, based on the maximum distance of 707 km for the combined upper Rift Valley and Eastern regions of northern Kenya (Journel & Huijbregts, 1978; Crawley, 2013).

**Figure 1.**
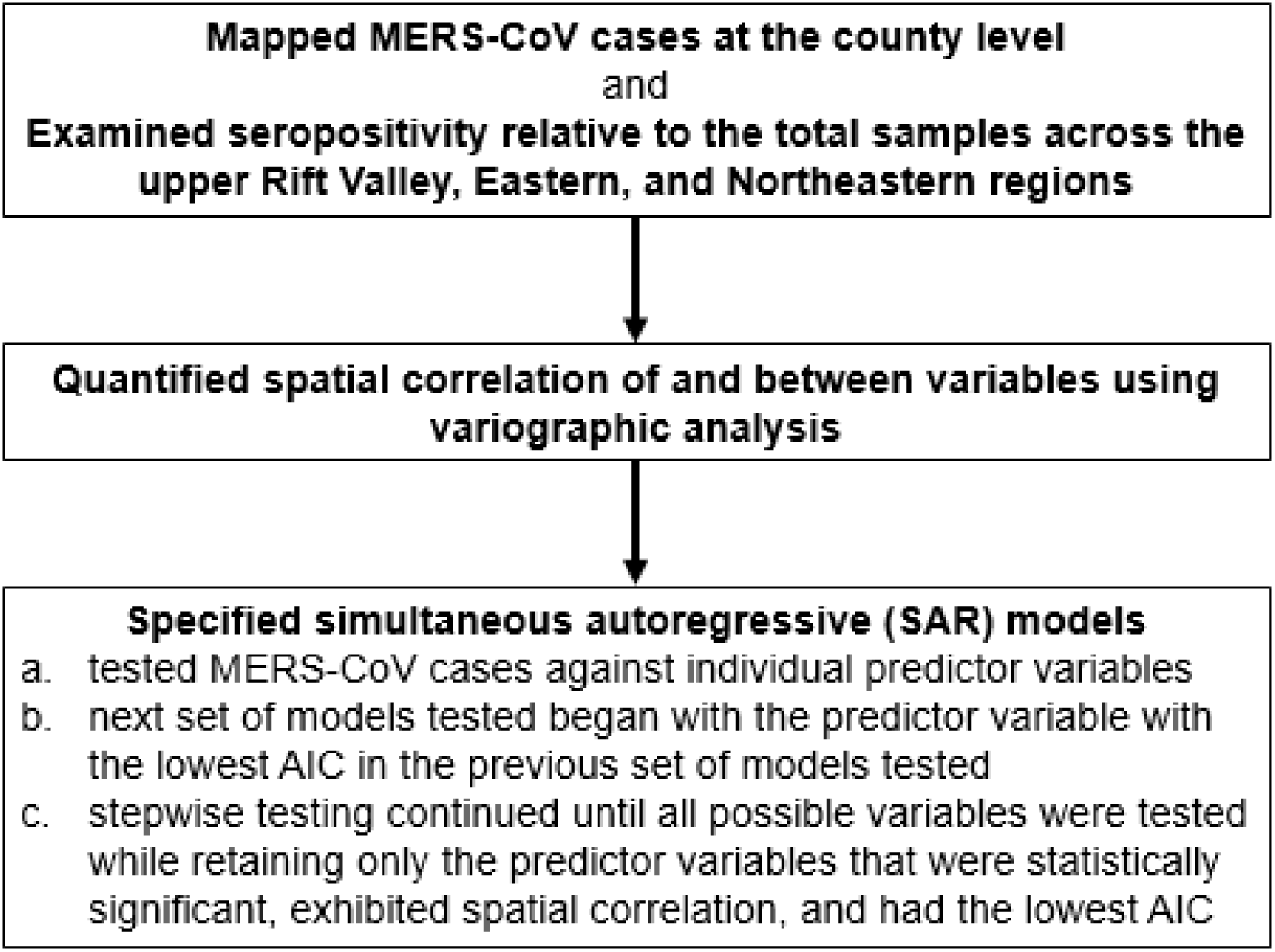
Summary process of spatial modeling and analysis of MERS-CoV and climate, agriculture, and socio-demographic factors related to agro-pastoralism that can influence the prevalence. The initial step involved mapping MERS-CoV cases and examining relative seropositivity across northern parts of regions across Kenya. The next step involved quantifying spatial correlation between the variables. Finally, simultaneous autoregressive models were tested.

After confirming spatial correlation among the MERS-CoV cases and socio-environmental variables, we specified simultaneous autoregressive (SAR) models, a statistical method that augments linear regression models with an additional term to account for the spatial correlation structure in a dataset (Kissling & Carl, 2008). To include the spatial correlation structure of our dataset into the SAR models, we defined neighbors among the northern and southern parts of and between each county based on shared borders, and created a spatially weighted matrix. Using shared borders to define neighbors, rather than including counties beyond those with shared borders, allowed us to account for spatial correlation if it diminished over an increasing distance. We weighted each county’s neighbor equally, such that the weights of all neighbors of a sub-county summed to one. Equation 1 shows the general SAR model in matrix form that includes the spatial structure of our dataset.

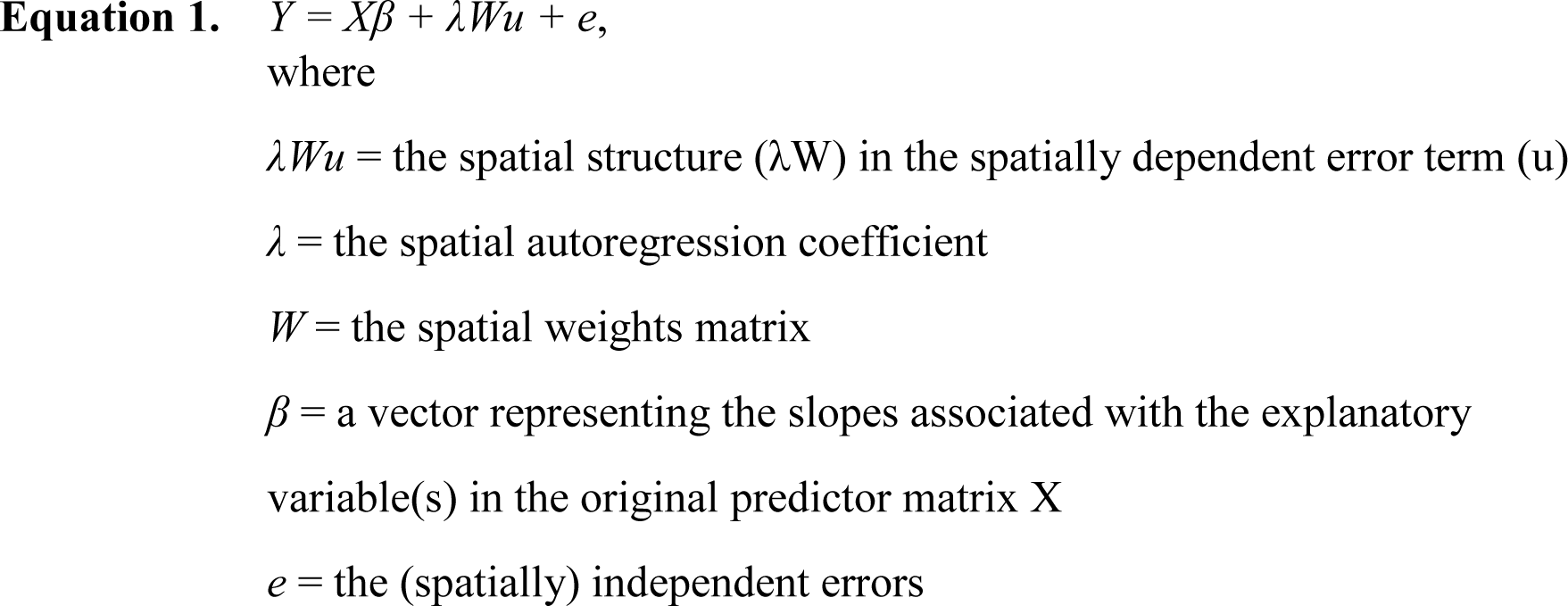

Our analyses involved testing SAR models in a stepwise process. First, we tested MERS-CoV cases against individual predictor variables. The predictor variables that were statistically significant and spatial correlated with MERS-CoV cases were retained for further testing in the next set of models. The next set of models tested began with the predictor variable with the lowest AIC in the previous set of models tested. Each of the other statistically significant predictor variables from the previously tested set of models were then individually tested in the new best model at that point in the process. The stepwise testing continued until all possible variables were tested while retaining only the predictor variables that were statistically significant, exhibited spatial correlation, and had the lowest AIC. We evaluated and compared the SAR models relative to each other using the p-value (with at least a 90% level) of the likelihood ratio test where a model with no spatial correlation (i.e., *λ* = 0) is compared to the fitted model with a non-zero spatial correlation parameter (Kissling & Carl, 2008). Ultimately, we used the Akaike information criterion (AIC) to choose the best performing model.

## 3. Results

The total number of samples and seropositive cases between 1992 through 2018 in the upper Rift Valley (Turkana, Baringo, W. Pokot, Laikipia) (n=2229, pos.= 914) had a 41% positivity, in the upper Eastern region (Marsabit, Isiolo, Samburu) (n=2486, pos.=1876) had a 75% positivity, and in the upper North Eastern region (Mandera, Wajir) (n=3506, pos.=1983) had a 57% positivity (Figure 2). The total number of samples and seropositive cases between 2016 through 2018 in the upper Rift Valley (n=935, pos.= 414) had a 51% positivity, and the upper Eastern region (n=1964, pos.=1510) had a 77% positivity. Spatial correlation of MERS-CoV extended a distance of roughly 400 kilometers across the upper Rift Valley and Eastern regions between 2016 and 2018.

**Figure 2.**
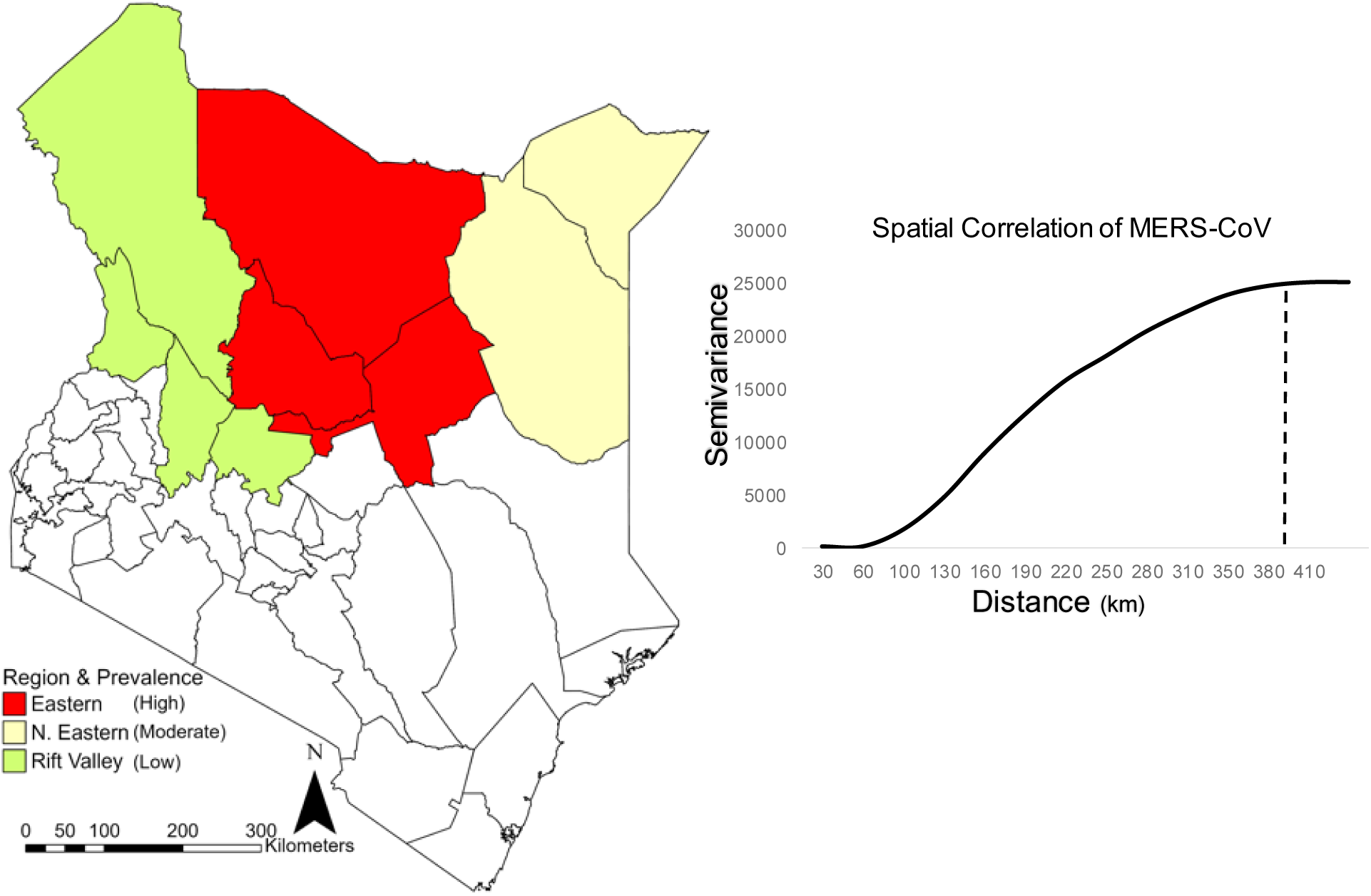
Geographic distribution and spatial correlation of MERS-CoV across northern regions of Kenya. The upper Rift Valley (counties of Turkana, Baringo, W. Pokot, Laikipia) had an average seropositivity of 41% between 1992 and 2018, and an average seropositivity of 51% between 2016 and 2018. The upper Eastern region (counties Marsabit, Isiolo, Samburu) had an average seropositivity of 75% between 1992 and 2018, and an average seropositivity of 77% between 2016 and 2018. The upper Northeastern region (counties of Mandera and Wajir) had a seropositivity of 57% in 2008. The spatial autocorrelation of MERS-CoV extends a distance of approximately 400 km in 2016 to 2018 across the upper Rift Valley and Eastern regions of northern Kenya.

The climate, agriculture, and socio-demographic variables (Appendix C) that were individually spatially correlated with MERS-CoV cases between 2016 and 2018 and statistically significant according to the p-value were included in the SAR models. While all of the six spatial covariates included in the SAR models did not achieve p < 0.10, they were deemed close to the cutoff and important theoretically to assess the spatial relationship (Table 2). According to the AIC, the spatial relationship of ethno-religious camel practices with MERS-CoV cases was significantly better than the other independent variables. Specifically, the p-value (p=0.015) was below the significance threshold of p < 0.05 and the AIC was statistically different from all the other individually tested independent variables with a -24.92 change in the AIC from the weakest performing SAR model, which was the spatial relationship of human population with MERS-CoV cases.

**Table 2.** SAR model results for initial models that tested variables in relation to MERS-CoV in dromedary camels across upper Rift Valley and Eastern regions of Kenya

| Model | Variables | Coefficient estimate | Standard error | Autoregressive coefficient ($\lambda$) | P value | AIC and $\Delta$ AIC |
| --- | --- | --- | --- | --- | --- | --- |
|  | Response Explanatory |  |  |  |  |  |
|  | MERS-CoV Prevalence in Camels |  |  |  |  |  |
| 1 | (Intercept)<br>Human Population | 119.64<br>-0.04 | 89.97<br>0.30 | 0.57 | 0.121 | 179.5 |
| 2 | (Intercept)<br>Ethnic Diversity | 100.19<br>2.34 | 114.44<br>18.76 | 0.58 | 0.079 | -0.01 |
| 3 | (Intercept)<br>Agropastoral Activities | 98.99<br>0.43 | 82.71<br>1.55 | 0.59 | 0.075 | -0.07 |
| 4 | (Intercept)<br>Agriculture Land | 171.70<br>-0.001 | 85.60<br>0.001 | 0.59 | 0.061 | -1.14 |
| 5 | (Intercept)<br>Avg. Air Temperature | -506.40<br>8.66 | 253.12<br>3.47 | 0.55 | 0.110 | -5.14 |
| 6 | (Intercept)<br>Ethno-religious Camel Practices | -37.23<br>3.33 | 45.24<br>0.39 | 0.74 | 0.015 | -24.92 |

The multi-variate SAR models that were statistically significant and spatially correlated with MERS-CoV cases are shown in Table 3, while all multi-variate SAR models that were tested are shown in Appendix D through Appendix H. The first synergistic effect appeared with the inclusion of agricultural land (model 9), which was statistically different from ethno-religious camel practices alone (model 6) based on the AIC, as well as spatially correlated with MERS-CoV cases. The SAR model improved, according to the AIC, and a broader synergistic effect demonstrated when ethnic diversity and agro-pastoral activities were included (model 19).

**Table 3.** Results of the relevant multi-variate SAR models that tested variables in relation to MERS-CoV in dromedary camels across upper Rift Valley and Eastern regions of Kenya

| Model | Variables | Coefficient estimate | Standard error | Autoregressive coefficient (λ) | P value | AIC |
| --- | --- | --- | --- | --- | --- | --- |
|  | Response |  |  |  |  |  |
|  | Explanatory |  |  |  |  |  |
| MERS-CoV Prevalence in Camels |  |  |  |  |  |  |
| 9 | (Intercept) | 22.51 | 38.07 | 0.74 | 0.029 | 147.89 |
|  | Ethno-religious Camel Practices | 3.31 | 0.29 |  |  |  |
|  | Agriculture Land | -0.0006 | 0.0002 |  |  |  |
| 19 | (Intercept) | -158.76 | 45.54 | 0.74 | 0.046 | 138.37 |
|  | Ethno-religious Camel Practices | 4.38 | 0.29 |  |  |  |
|  | Agriculture Land | 0.0007 | 0.0003 |  |  |  |
|  | Ethnic Diversity | 21.74 | 4.95 |  |  |  |
|  | Agropastoral Activities | -3.47 | 0.80 |  |  |  |
| 23 | (Intercept) | -501.65 | 65.35 | -1.39 | 0.008 | 95.64 |
|  | Ethno-religious Camel Practices | 3.09 | 0.16 |  |  |  |
|  | Agriculture Land | 0.0001 | 0.00005 |  |  |  |
|  | Ethnic Diversity | 12.08 | 0.82 |  |  |  |
|  | Agropastoral | -2.22 | 0.613 |  |  |  |
|  | Average Temperature | 7.57 | 1.10 |  |  |  |
|  | Human Population | -0.34 | 0.08 |  |  |  |

However, the best performing SAR model according to the AIC, and the most complex synergistic effect was when all of the six predictors (model 23) were included, and were spatially correlated with MERS-CoV cases and statistically significant. Thus, ethno-religious camel practices and agriculture land were the two initially interacting variables of importance. Next, ethnic diversity and agro-pastoral activities combined with the previous two independent variables were the next set of interacting variables of importance. Finally, average air temperature and human population combined with the previous four independent variables were the final set of interacting variables of importance.

## 4. Discussion

This study was conducted to collate data from disparate studies and datasets to provide greater insight to the prediction of MERS-CoV infections that may be facing Kenya, and regions with similar geographies across the world. Most often, the occurrence of infectious diseases has been studied independently, overlooking the potential combined effects of climate change, agricultural land use, and human-environment interaction. Here, we comprehensively and spatially evaluated the influence of various components that may contribute to the emergence of MERS-CoV throughout the northern regions of Kenya. This study highlighted the benefit of bringing together diverse areas of data and research to spatially inform infectious disease risk.

The geographical variations of serological evidence of MERS-CoV in dromedary camels and spatial correlations with the social and environmental variables may be partly attributed to the dominant ethnic groups in the different regions and their particular agro-pastoral management practices. Our results showed low MERS-CoV seropositivity in the Rift Valley region where tribal communities practice a more diversified form of livestock management with camels reared alongside other livestock, such as cattle, sheep, and goats (Iiyama et al., 2008).

Particularly, the Turkana communities and the Pokot have adapted livestock diversification as part of a long-term adaptation strategy to manage drought and diseases (Opiyo et al., 2015), and tend to acquire cattle and camels through cultural practices, such as dowry rather than directly from markets (De Vries et al., 2006). Thus, tribal communities, such as Pokot, Maasai or Turkana have a limited dependence on camels for their economic activity that can result in lower seropositivity of MER-CoV compared to other regions (Deem et al., 2015).

Tribal communities, such as the Somali, Gabra, and Borana in the eastern and northeastern regions of Kenya primarily focus on camels as a pastoral livelihood strategy due to the camel’s adaptability to arid environments and ability to withstand droughts (Ngere et al., 2020). Also, the Somali primarily inhabit the Northeastern region with high regard for camels for their annual religious practices and ritual migrations (Watson, 2010), and a greater number of cultural practices and traditions associated with livestock rearing, trade, and consumption in tribal communities (Dan et al., 2021; Kagunyu et al., 2018). Further, the tribal communities in both the Eastern and Northeastern regions tend to migrate to different areas in search of fresh water and pastures, and restock their camels through direct markets, which can lead to higher prevalence rates of MERS-CoV (De Vries et al., 2006; (Hughes & Anderson, 2020).

Recent findings have shown that the expanding arid regions of Kenya have reduced agricultural land use and increased the reliance on pastoralism, and particularly camel rearing (Lawrence et al., 2023a). Also, the increasing trend in air temperatures has resulted in decreased rainfed agricultural production, and further leading to increased pastoralism (Lawrence et al., 2023b). As a consequence, these effects of climate change may continue to pose a major challenge in creating favorable conditions for the emergence and transmission of viral zoonotic diseases resulting in the observed high seropositivity rates of MERS-CoV. Similar results of spatial clustering of zoonotic diseases have been studied in different agro-ecological climate zones and sociodemographics of Kenya to identify potential disease hotspots of anthrax (Nderitu et al., 2021). Overall, these findings are significant as shifting climatic zones continue to impact agricultural systems while zoonotic diseases like MERS-CoV pose a major risk to communities living in arid regions of Kenya warranting immediate interventions and continued surveillance.

Due to these climatic and agricultural shifts, there is an urgent need to predict where and under what conditions would viral transmission of MERS-CoV occur. These findings suggest that continued monitoring of camels, humans, and their spatial patterns will be an integral component of informing prediction models. This study relied on previously collected data that differed in methods and specificity of data, which also assumes equal probability of detecting seropositivity. Further, the county-level finding in this study can vary at smaller more local spatial scales. Thus, results are limited in power and yet, the broader spatial patterns observed and insights that this type of study can provide are important in the planning and implementation of prediction modeling, as well as location-based risk management.

## 5. Conclusion

Climate and agricultural land-use change have increased the likelihood of infectious disease emergence and transmissions, but these drivers are often examined separately as synergistic effects are ignored. Further, seldom are the influence of climate and agricultural land use on emerging infectious diseases examined in a spatially explicit way at regional scales. Our objective in this study was to spatially examine the climate, agriculture, and socio-demographic factors related to agro-pastoralism that can influence the prevalence of MERS-CoV in dromedary camels across northern Kenya. Overall, this study can provide important insights in the planning and implementation of prediction modeling, as well as location-based risk management.

## Data Availability

All data produced in the present study are available upon reasonable request to the authors

## Appendix A. Kenya Regional and County Maps. Kenya comprises 8 regions (a) and 47 counties (b)

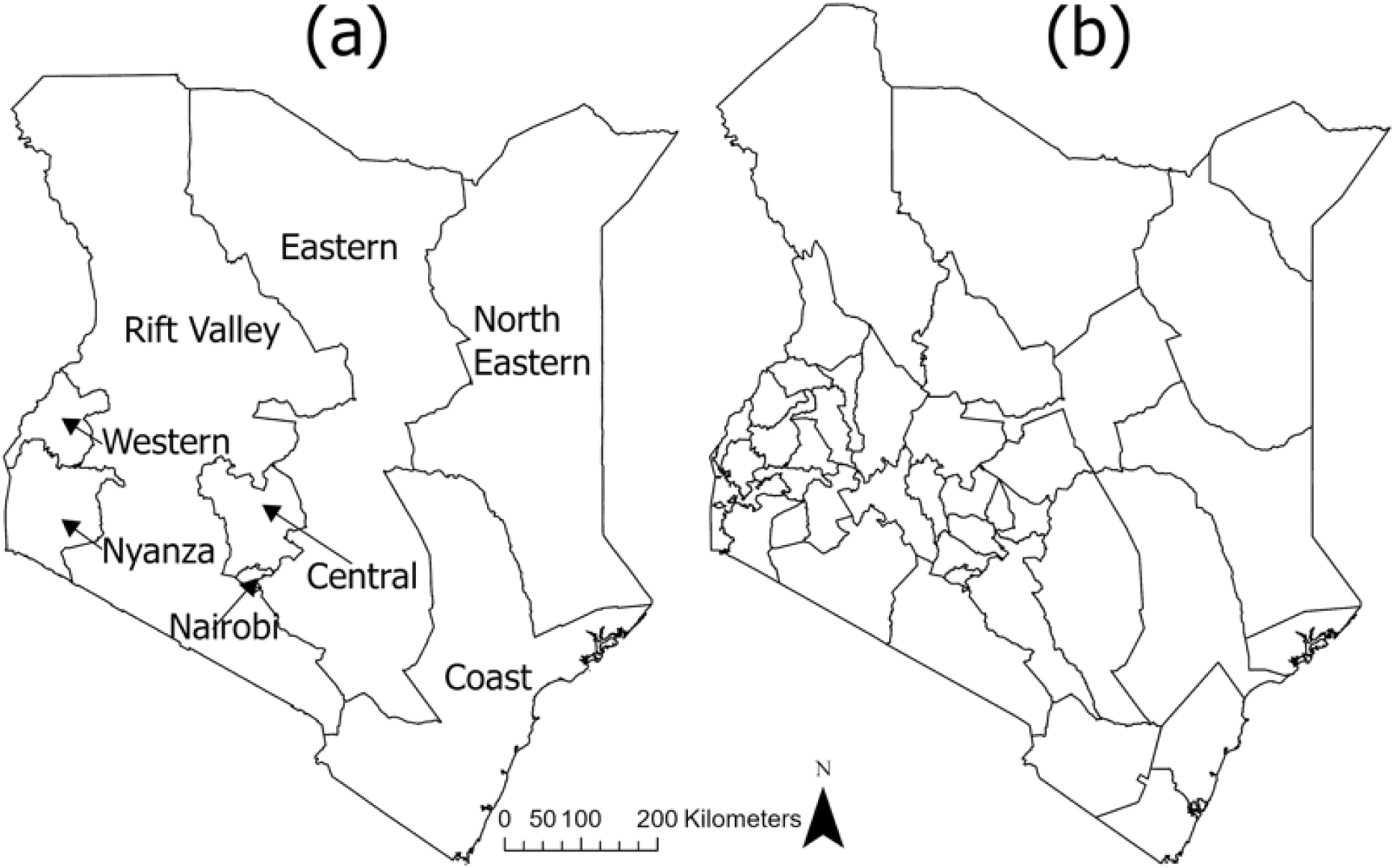

## Appendix B. Previous MERS-CoV studies in Kenya that report seropositivity

## Appendix C. Independent variables tested in spatial statistical models to predict MERS-CoV Category

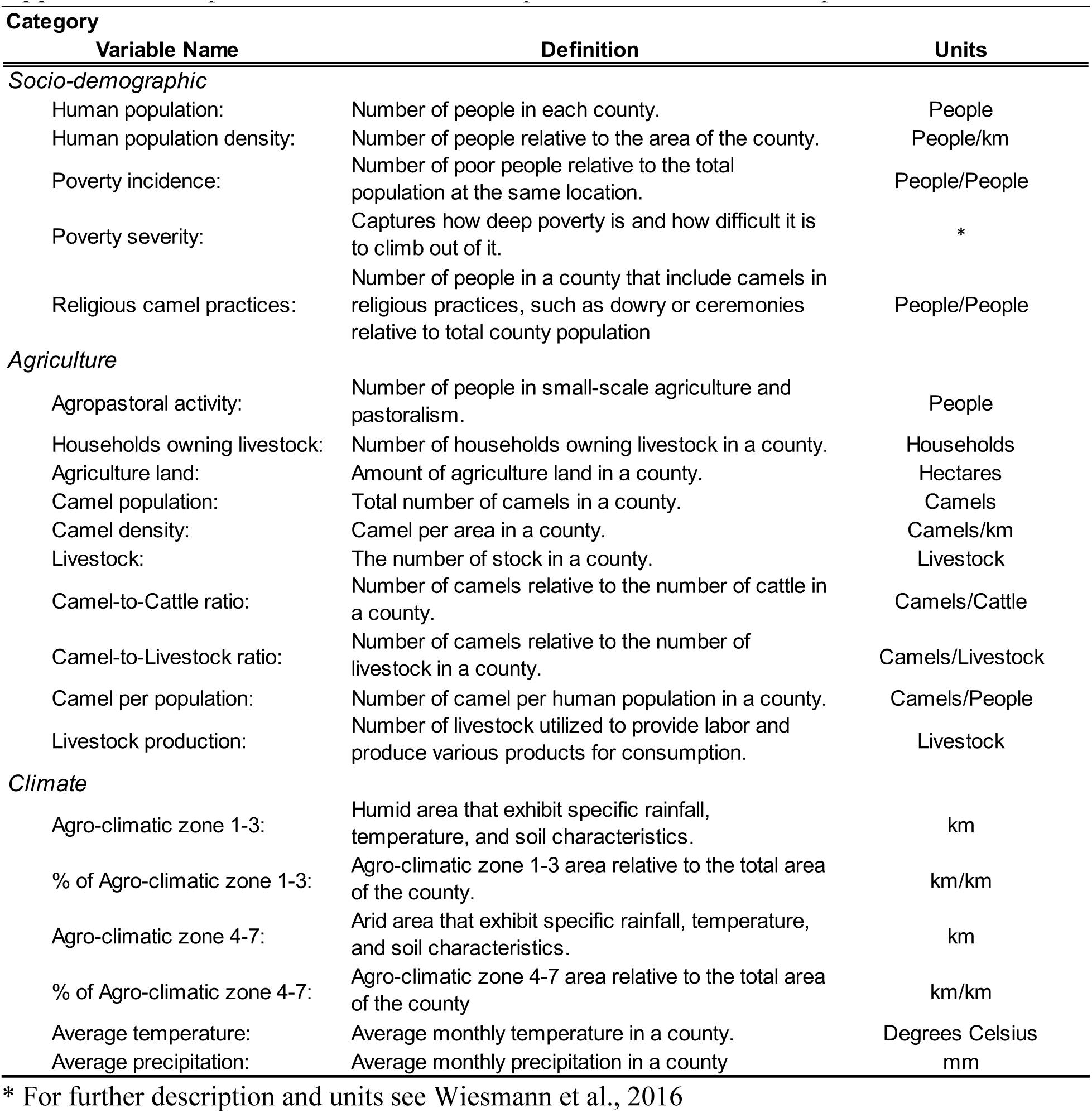

## Appendix D Second set of models that tested variables in relation to MERS-CoV in dromedary camels across upper Rift Valley and Eastern regions of Kenya

| Model | Variables | | Coefficient estimate | Standard error | Autoregressive | | AIC and $\Delta$ AIC |
| --- | --- | --- | --- | --- | --- | --- | --- |
| | Response | Explanatory | | | coefficient ($\lambda$) | P value | |
| MERS-CoV Prevalence in Camels |  |  |  |  |  |  |  |
| 6 |  | (Intercept) | -37.23 | 45.24 | 0.74 | 0.015 | 154.58 |
|  |  | Ethno-religious Camel Practices | 3.33 | 0.39 |  |  |  |
| 7 |  | (Intercept) | 194.17 | 146.34 | 0.74 | 0.019 | -0.49 |
|  |  | Ethno-religious Camel Practices | 3.95 | 0.52 |  |  |  |
|  |  | Avg. Air Temperature | -3.63 | 2.20 |  |  |  |
| 8 |  | (Intercept) | -106.39 | 54.54 | 0.74 | 0.018 | -1.30 |
|  |  | Ethno-religious Camel Practices | 3.43 | 0.35 |  |  |  |
|  |  | Ethnic Diversity | 12.93 | 6.70 |  |  |  |
| 9 |  | (Intercept) | 22.51 | 38.07 | 0.74 | 0.029 | -6.69 |
|  |  | Ethno-religious Camel Practices | 3.31 | 0.29 |  |  |  |
|  |  | Agriculture Land | -0.0006 | 0.0001 |  |  |  |
| 10 |  | (Intercept) | -6.99 | 24.64 | 0.56 | 0.240 | -7.77 |
|  |  | Ethno-religious Camel Practices | 3.87 | 0.31 |  |  |  |
|  |  | Agropastoral | -1.91 | 0.49 |  |  |  |
| 11 |  | (Intercept) | 25.60 | 14.53 | -0.27 | 0.634 | -15.47 |
|  |  | Ethno-religious Camel Practices | 4.09 | 0.21 |  |  |  |
|  |  | Human Population | -0.48 | 0.05 |  |  |  |

## Appendix E Third set of models that tested variables in relation to MERS-CoV in dromedary camels across upper Rift Valley and Eastern regions of Kenya

| Model | Variables | | Coefficient estimate | Standard error | Autoregressive | | AIC and $\Delta$ AIC |
| --- | --- | --- | --- | --- | --- | --- | --- |
|  | Response | Explanatory |  |  | coefficient | P value |  |
| | | | | | ($\lambda$) | | |
| MERS-CoV Prevalence in Camels |  |  |  |  |  |  |  |
| 9 | (Intercept) |  | 22.51 | 38.07 | 0.74 | 0.029 | 147.89 |
|  | Ethno-religious Camel Practices |  | 3.31 | 0.29 |  |  |  |
|  | Agriculture Land |  | -0.0006 | 0.0002 |  |  |  |
| 12 | (Intercept) |  | -217.65 | 164.42 | 0.72 | 0.046 | -0.05 |
|  | Ethno-religious Camel Practices |  | 2.57 | 0.57 |  |  |  |
|  | Agriculture Land |  | -0.0009 | 0.00027 |  |  |  |
|  | Average Temperature |  | 4.30 | 2.89 |  |  |  |
| 13 | (Intercept) |  | -22.48 | 53.66 | 0.79 | 0.013 | 0.40 |
|  | Ethno-religious Camel Practices |  | 3.39 | 0.27 |  |  |  |
|  | Agriculture Land |  | -0.0005 | 0.00016 |  |  |  |
|  | Ethnic Diversity |  | 7.21 | 5.44 |  |  |  |
| 14 | (Intercept) |  | 5.99 | 31.77 | 0.64 | 0.18 | 0.46 |
|  | Ethno-religious Camel Practices |  | 3.68 | 0.36 |  |  |  |
|  | Agriculture Land |  | -0.0002 | 0.0003 |  |  |  |
|  | Agropastoral |  | -1.260 | 0.923 |  |  |  |
| 15 | (Intercept) |  | 25.33 | 14.54 | -0.27 | 0.618 | -6.83 |
|  | Ethno-religious Camel Practices |  | 4.09 | 0.21 |  |  |  |
|  | Agriculture Land |  | 0.00004 | 0.00017 |  |  |  |
|  | Human Population |  | -0.50 | 0.09 |  |  |  |

## Appendix F Fourth set of models that tested variables in relation to MERS-CoV in dromedary camels across upper Rift Valley and Eastern regions of Kenya

| Model | Variables | | Coefficient estimate | Standard error | Autoregressive | | AIC and $\Delta$ AIC |
| --- | --- | --- | --- | --- | --- | --- | --- |
| | Response | Explanatory | | | coefficient ($\lambda$) | P value | |
| MERS-CoV Prevalence in Camels |  |  |  |  |  |  |  |
| 12 | (Intercept) |  | -217.65 | 164.42 | 0.72 | 0.046 | 147.84 |
|  | Ethno-religious Camel Practices |  | 2.58 | 0.57 |  |  |  |
|  | Agriculture Land |  | -0.0009 | 0.0003 |  |  |  |
|  | Average Temperature |  | 4.30 | 2.89 |  |  |  |
| 16 | (Intercept) |  | -171.47 | 187.71 | 0.75 | 0.051 | 1.83 |
|  | Ethno-religious Camel Practices |  | 2.79 | 0.75 |  |  |  |
|  | Agriculture Land |  | -0.0008 | 0.0004 |  |  |  |
|  | Average Temperature |  | 3.13 | 3.82 |  |  |  |
|  | Ethnic Diversity |  | 3.14 | 7.29 |  |  |  |
| 17 | (Intercept) |  | -784.30 | 83.87 | -0.52 | 0.318 | -20.99 |
|  | Ethno-religious Camel Practices |  | 1.93 | 0.31 |  |  |  |
|  | Agriculture Land |  | -0.0005 | 0.00012 |  |  |  |
|  | Average Temperature |  | 13.78 | 1.50 |  |  |  |
|  | Agropastoral |  | -4.13 | 0.39 |  |  |  |
| 18 | (Intercept) |  | -407.43 | 77.45 | -0.27 | 0.599 | -21.30 |
|  | Ethno-religious Camel Practices |  | 2.54 | 0.30 |  |  |  |
|  | Agriculture Land |  | -0.00044 | 0.00013 |  |  |  |
|  | Average Temperature |  | 7.80 | 1.39 |  |  |  |
|  | Human Population |  | -0.52 | 0.05 |  |  |  |

## Appendix G Fifth set of models that tested variables in relation to MERS-CoV in dromedary camels across upper Rift Valley and Eastern regions of Kenya

| Model | Variables | | Coefficient estimate | Standard error | Autoregressive | | AIC and $\Delta$ AIC |
| --- | --- | --- | --- | --- | --- | --- | --- |
|  | Response | Explanatory |  |  | coefficient | P value |  |
| | | | | | ($\lambda$) | | |
| MERS-CoV Prevalence in Camels |  |  |  |  |  |  |  |
| 13 |  | (Intercept) | -22.48 | 53.66 | 0.79 | 0.013 | 148.29 |
|  |  | Ethno-religious Camel Practices | 3.39 | 0.27 |  |  |  |
|  |  | Agriculture Land | -0.0005 | 0.00016 |  |  |  |
|  |  | Ethnic Diversity | 7.21 | 5.44 |  |  |  |
| 19 |  | (Intercept) | -158.76 | 45.54 | 0.74 | 0.046 | -9.92 |
|  |  | Ethno-religious Camel Practices | 4.38 | 0.29 |  |  |  |
|  |  | Agriculture Land | 0.0007 | 0.0003 |  |  |  |
|  |  | Ethnic Diversity | 21.74 | 4.95 |  |  |  |
|  |  | Agropastoral | -3.47 | 0.80 |  |  |  |
| 20 |  | (Intercept) | -97.90 | 16.57 | -0.54 | 0.384 | -28.77 |
|  |  | Ethno-religious Camel Practices | 4.41 | 0.09 |  |  |  |
|  |  | Agriculture Land | 0.001 | 0.0001 |  |  |  |
|  |  | Ethnic Diversity | 17.95 | 2.27 |  |  |  |
|  |  | Human Population | -0.662 | 0.042 |  |  |  |

## Appendix H Sixth set of models that tested variables in relation to MERS-CoV in dromedary camels across upper Rift Valley and Eastern regions of Kenya

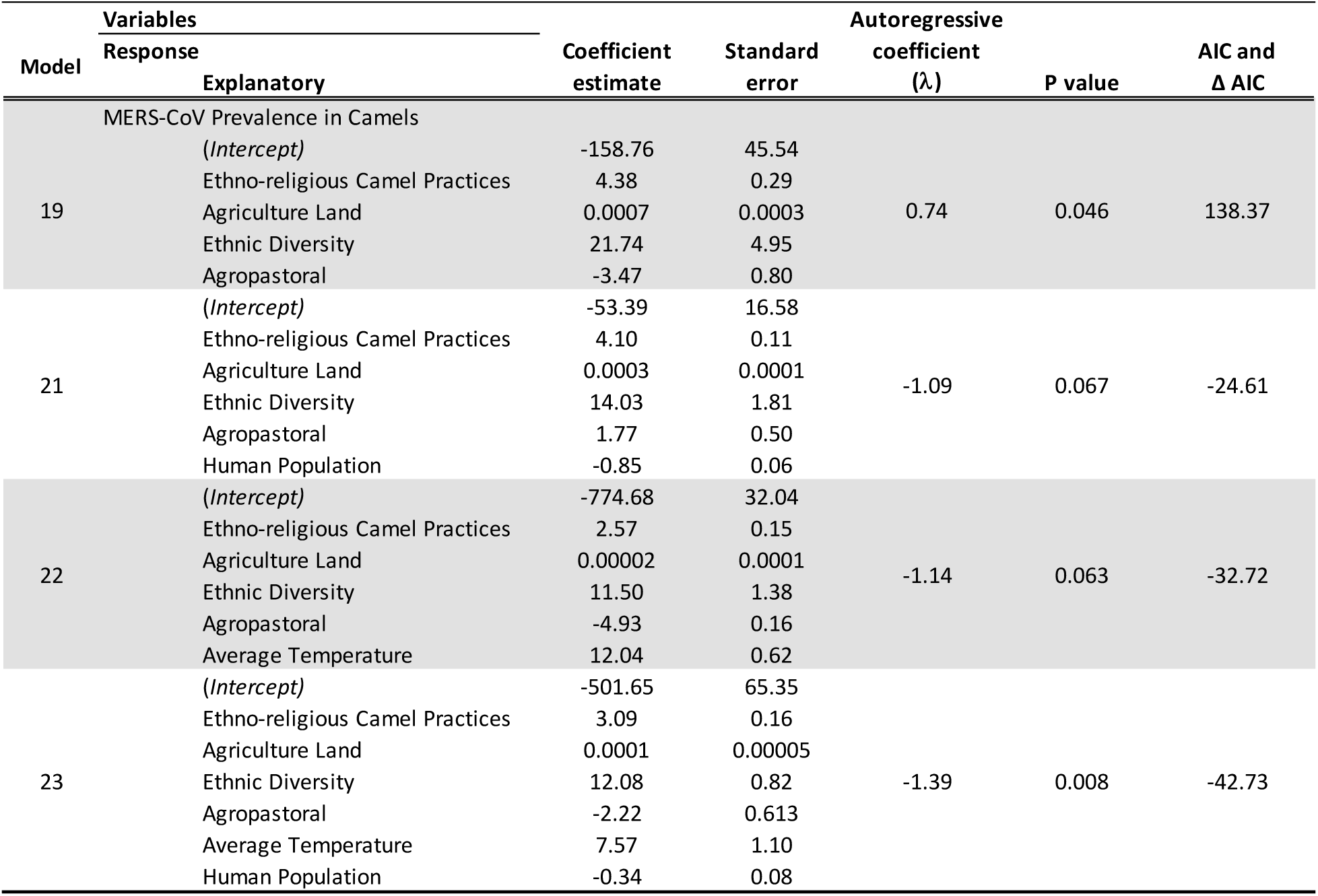

## Notes

### Competing Interest Statement

The authors have declared no competing interest.

### Funding Statement

This work was supported in part by the Taylor Geospatial Institute and a seed research grant from the Living Earth Collaborative at Washington University in St. Louis. Furthermore, this research was also supported in part by the UK Biotechnology and Biological Sciences Research Council, the Department for International Development, the Economic & Social Research Council, the Medical Research Council, the Natural Environment Research Council, and the Defense Science & Technology Laboratory, under the Zoonoses and Emerging Livestock Systems (ZELS) programme, grant reference BB/L019019/1. This study also received support from the CGIAR One Health initiative Protecting Human Health Through a One Health Approach, which was supported by contributors to the CGIAR Trust Fund (https://www.cgiar.org/funders/). We thank the University of Liverpool Open Access team for support of the CC-BY open access license for this article.

## References

1. Adney DR, van Doremalen N, Brown VR, et al. (2014) Replication and Shedding of MERS-CoV in Upper Respiratory Tract of Inoculated Dromedary Camels. Emerging Infectious Diseases 20:1999–2005; doi:10.3201/eid2012.141280

2. Aguanno R, ElIdrissi A, Elkholy AA, et al. (2018) MERS: Progress on the global response, remaining challenges and the way forward. Antiviral Research 159:35–44; doi:10.1016/j.antiviral.2018.09.002

3. Baker RE, Mahmud AS, Miller IF, Rajeev M, Rasambainarivo F, Rice BL, Takahashi S, Tatem AJ, Wagner CE, Wang L-F, Wesolowski A, Metcalf CJE (2022) Infectious disease in an era of global change. Nature Reviews Microbiology 20(4), Article 4; 10.1038/s41579-021-00639-z

4. Boitt MK, Mundia CN, Pellikka P (2014) Modelling the Impacts of Climate Change on Agro-Ecological Zones – a Case Study of Taita Hills, Kenya. Universal Journal of Geoscience, 8.

5. Brodie JF (2016) Synergistic effects of climate change and agricultural land use on mammals. Frontiers in Ecology and the Environment 14:20–26; 10.1002/16-0110.1

6. Bullock EL, Healey SP, Yang Z, Oduor P, Gorelick N, Omondi S, Ouko E, Cohen WB (2021) Three Decades of Land Cover Change in East Africa. Land 10:150; 10.3390/land10020150

7. Cecchi G, Wint W, Shaw A, Marletta A, Mattioli R, Robinson T (2010) Geographic distribution and environmental characterization of livestock production systems in Eastern Africa. Agriculture, Ecosystems & Environment 135:98–110; 10.1016/j.agee.2009.08.011

8. Dan JM, Mateus J, Kato Y, Hastie KM, Yu ED, Faliti CE, Grifoni A, Ramirez SI, Haupt S, Frazier A, Nakao C, Rayaprolu V, Rawlings SA, Peters B, Krammer F, Simon V, Saphire EO, Smith DM, Weiskopf D, et al. (2021) Immunological memory to SARS-CoV-2 assessed for up to 8 months after infection. Science 371(6529), eabf4063; 10.1126/science.abf4063

9. De Vries D, Leslie PW, McCabe JT (2006) Livestock Acquisitions Dynamics in Nomadic Pastoralist Herd Demography: A Case Study Among Ngisonyoka Herders of South Turkana, Kenya. Human Ecology 34:1–25; 10.1007/s10745-005-9000-2

10. Deem SL, Fèvre EM, Kinnaird M, Browne AS, Muloi D, Godeke G-J, Koopmans M, Reusken CB (2015) Serological Evidence of MERS-CoV Antibodies in Dromedary Camels (Camelus dromedaries) in Laikipia County, Kenya. PLOS ONE 10:e0140125; 10.1371/journal.pone.0140125

11. Duygu F, Sari T, Kaya T, Tavsan O, Naci M (2018) THE RELATIONSHIP BETWEEN CRIMEAN-CONGO HEMORRHAGIC FEVER AND CLIMATE: DOES CLIMATE AFFECT THE NUMBER OF PATIENTS? Acta Clinica Croatica 57:443–448; 10.20471/acc.2018.57.03.06

12. Fanelli A, Buonavoglia D (2021) Risk of Crimean Congo haemorrhagic fever virus (CCHFV) introduction and spread in CCHF-free countries in southern and Western Europe: A semi-quantitative risk assessment. One Health 13; 10.1016/j.onehlt.2021.100290

13. Fenollar F, Mediannikov O (2018) Emerging infectious diseases in Africa in the 21st century. New Microbes and New Infections 26:S10–S18; 10.1016/j.nmni.2018.09.004

14. Gikonyo S, Kimani T, Matere J, Kimutai J, Kiambi SG, Bitek AO, Juma Ngeiywa KJZ, Makonnen YJ, Tripodi A, Morzaria S, Lubroth J, Rugalema G, Fasina FO (2018) Mapping Potential Amplification and Transmission Hotspots for MERS-CoV, Kenya. EcoHealth 15:372–387; 10.1007/s10393-018-1317-6

15. Heffernan C (2018) Climate change and multiple emerging infectious diseases. Veterinary Journal 234:43–47; 10.1016/j.tvjl.2017.12.021

16. Hughes EC, Anderson NE (2020) Zoonotic Pathogens of Dromedary Camels in Kenya: A Systematised Review. Veterinary Sciences 7(3), Article 3; 10.3390/vetsci7030103

17. Iiyama M, Kariuki P, Kristjanson P, Kaitibie S, Maitima J (2008) Livelihood diversification strategies, incomes and soil management strategies: A case study from Kerio Valley, Kenya. Journal of International Development 20:380–397; 10.1002/jid.1419

18. Kagunyu A, Lengarite M, Chemuluti J (2018) Analyzing the Social—Economic Issues Surrounding Camel Production in Northern Kenya. Environmental Research Journal 11:95– 102.

19. Kissling WD, Carl G (2008) Spatial autocorrelation and the selection of simultaneous autoregressive models. Global Ecology and Biogeography 17:59–71; 10.1111/j.1466-8238.2007.00334.x

20. Koeva M, Stöcker C, Crommelinck S, Ho S, Chipofya M, Sahib J, Bennett R, Zevenbergen J, Vosselman G, Lemmen C, Crompvoets J, Buntinx I, Wayumba G, Wayumba R, Odwe PO, Osewe GT, Chika B, Pattyn V (2020) Innovative Remote Sensing Methodologies for Kenyan Land Tenure Mapping. Remote Sensing 12(2), Article 2; 10.3390/rs12020273

21. Lawrence TJ, Vilbig JM, Kangogo G, Fèvre EM, Deem SL, Gluecks I, Sagan V, Shacham E (2023a) Shifting climate zones and expanding tropical and arid climate regions across Kenya (1980–2020). Regional Environmental Change 23:59; 10.1007/s10113-023-02055-w

22. Lawrence TJ, Vilbig JM, Kangogo G, Fèvre EM, Deem SL, Gluecks I, Sagan V, Shacham E (2023b) Spatial changes to climatic suitability and availability of agropastoral farming systems across Kenya (1980–2020). Outlook on Agriculture 52:186–199; 10.1177/00307270231176577

23. Lee-Cruz L, Lenormand M, Cappelle J, Caron A, Nys HD, Peeters M, Bourgarel M, Roger F, Tran A (2021) Mapping of Ebola virus spillover: Suitability and seasonal variability at the landscape scale. PLOS Neglected Tropical Diseases 15:e0009683; 10.1371/journal.pntd.0009683

24. Muturi M, Mwatondo A, Nijhof AM, Akoko J, Nyamota R, Makori A, Nyamai M, Nthiwa D, Wambua L, Roesel K, Thumbi SM, Bett B (2023) Ecological and subject-level drivers of interepidemic Rift Valley fever virus exposure in humans and livestock in Northern Kenya. Scientific Reports 13:15342; 10.1038/s41598-023-42596-y

25. Nderitu LM, Gachohi J, Otieno F, Mogoa EG, Muturi M, Mwatondo A, Osoro EM, Ngere I, Munyua PM, Oyas H, Njagi O, Lofgren E, Marsh T, Widdowson MA, Bett B, Njenga MK (2021) Spatial clustering of livestock Anthrax events associated with agro-ecological zones in Kenya, 1957–2017. BMC Infectious Diseases 21:191; 10.1186/s12879-021-05871-9

26. Ngere I, Munyua P, Harcourt J, Hunsperger E, Thornburg N, Muturi M, Osoro E, Gachohi J, Bodha B, Okotu B, Oyugi J, Jaoko W, Mwatondo A, Njenga K, Widdowson MA (2020) High MERS-CoV seropositivity associated with camel herd profile, husbandry practices and household socio-demographic characteristics in Northern Kenya. Epidemiology and Infection 148:e292; 10.1017/S0950268820002939

27. Ngugi RK, Nyariki DM (2005) Rural livelihoods in the arid and semi-arid environments of Kenya: Sustainable alternatives and challenges. Agriculture and Human Values 22:65–71; 10.1007/s10460-004-7231-2

28. Opiyo F, Wasonga O, Nyangito M, Schilling J, Munang R (2015) Drought Adaptation and Coping Strategies Among the Turkana Pastoralists of Northern Kenya. International Journal of Disaster Risk Science 6:295–309; 10.1007/s13753-015-0063-4

29. Pandit PS, Anthony SJ, Goldstein T, Olival KJ, Doyle MM, Gardner NR, Bird B, Smith W, Wolking D, Gilardi K, Monagin C, Kelly T, Uhart MM, Epstein JH, Machalaba C, Rostal M K, Dawson P, Hagan E, Sullivan A, et al. (2022) Predicting the potential for zoonotic transmission and host associations for novel viruses. Communications Biology 5:844; 10.1038/s42003-022-03797-9

30. Recha CW (2019) REGIONAL VARIATIONS AND CONDITIONS FOR AGRICULTURE IN KENYA. Current Politics and Economics of Africa 12: 45.

31. Rohr JR, Barrett CB, Civitello DJ, Craft ME, Delius B, DeLeo GA, Hudson PJ, Jouanard N, Nguyen KH, Ostfeld RS, Remais JV, Riveau G, Sokolow SH, Tilman D (2019) Emerging human infectious diseases and the links to global food production. Nature Sustainability 2: 445–456; 10.1038/s41893-019-0293-3

32. Ryan SJ, Carlson CJ, Mordecai EA, Johnson LR (2019) Global expansion and redistribution of Aedes-borne virus transmission risk with climate change. PLoS Neglected Tropical Diseases 13:e0007213; 10.1371/journal.pntd.0007213

33. Semenza JC, Rocklöv J, Ebi KL (2022) Climate Change and Cascading Risks from Infectious Disease. Infectious Diseases and Therapy 11:1371–1390; 10.1007/s40121-022-00647-3

34. Serdeczny O, Adams S, Baarsch F, Coumou D, Robinson A, Hare W, Schaeffer M, Perrette M, Reinhardt J (2017) Climate change impacts in Sub-Saharan Africa: From physical changes to their social repercussions. Regional Environmental Change 17:1585–1600; 10.1007/s10113-015-0910-2

35. Xiao Y, Beier JC, Cantrell RS, Cosner C, DeAngelis DL, Ruan S (2015) Modelling the effects of seasonality and socioeconomic impact on the transmission of rift valley Fever virus. PLoS Neglected Tropical Diseases 9:e3388; 10.1371/journal.pntd.0003388

36. Saqib M, Sieberg A, Hussain MH, et al. (2017) Serologic Evidence for MERS-CoV Infection in Dromedary Camels, Punjab, Pakistan, 2012–2015. Emerging Infectious Diseases 23:550–551; doi:10.3201/eid2303.161285

37. WC-Kenya (2023) https://tcktcktck.org/kenya#climatic

38. WHO (2018) Geneva, Switzerland: World Health Organization. WHO MERS-CoV Global Summary and Assessment of Risk, August 2018 (WHO/MERS/RA/August18). Published online 2018. Licence: CC BY-NC-SA 3.0 IGO.

39. WHO (2019) Geneva, Switzerland: World Health Organization. WHO MERS-CoV Global Summary and Assessment of Risk, July 2019 (WHO/MERS/RA/19.1). Published July 2019. Accessed March 23, 2021. Licence: CC BY-NC-SA 3.0 IGO.

40. Wiesmann U, Kiteme B, Mwangi Z (2016) Socio-Economic Atlas of Kenya: Depicting the National Population Census by County and Sub-Location. Second, revised edition. KNBS, Nairobi. CETRAD, Nanyuki. CDE, Bern; 10.7892/boris.83693

## References

1. Corman, V. M., Jores, J., Meyer, B., Younan, M., Liljander, A., Said, M. Y., Gluecks, I., Lattwein, E., Bosch, B.-J., Drexler, J. F., Bornstein, S., Drosten, C., & Müller, M. A. (2014). Antibodies against MERS Coronavirus in Dromedary Camels, Kenya, 1992–2013. Emerging Infectious Diseases, 20(8). 10.3201/eid2008.140596

2. Deem, S. L., Fèvre, E. M., Kinnaird, M., Browne, A. S., Muloi, D., Godeke, G.-J., Koopmans, M., & Reusken, C. B. (2015). Serological Evidence of MERS-CoV Antibodies in Dromedary Camels (Camelus dromedaries) in Laikipia County, Kenya. PLOS ONE, 10(10), e0140125. 10.1371/journal.pone.0140125

3. Gardner, E. G., Kiambi, S., Sitawa, R., Kelton, D., Kimutai, J., Poljak, Z., Tadesse, Z., Von Dobschuetz, S., Wiersma, L., & Greer, A. L. (2019). Force of infection of Middle East respiratory syndrome in dromedary camels in Kenya. Epidemiology and Infection, 147. 10.1017/s0950268819001663

4. Ngere, I., Munyua, P., Harcourt, J., Hunsperger, E., Thornburg, N., Muturi, M., Osoro, E., Gachohi, J., Bodha, B., Okotu, B., Oyugi, J., Jaoko, W., Mwatondo, A., Njenga, K., & Widdowson, M. A. (2020). High MERS-CoV seropositivity associated with camel herd profile, husbandry practices and household socio-demographic characteristics in Northern Kenya. Epidemiology and Infection, 148. 10.1017/s0950268820002939

5. Ommeh, S., Zhang, W., Zohaib, A., Chen, J., Zhang, H., Hu, B., Ge, X.-Y., Yang, X.-L., Masika, M., Obanda, V., Luo, Y., Li, S., Waruhiu, C., Li, B., Zhu, Y., Ouma, D., Odendo, V., Wang, L.-F., Anderson, D. E., … Shi, Z.-L. (2018). Genetic Evidence of Middle East Respiratory Syndrome Coronavirus (MERS-Cov) and Widespread Seroprevalence among Camels in Kenya. Virologica Sinica, 33(6), 484–492. 10.1007/s12250-018-0076-4

6. Sitawa, R., Folorunso, F., Obonyo, M., Apamaku, M., Kiambi, S., Gikonyo, S., Kiptiness, J., Njagi, O., Githinji, J., Ngoci, J., VonDobschuetz, S., Morzaria, S., Ihab, E., Gardner, E., Wiersma, L., & Makonnen, Y. (2020). Risk factors for serological evidence of MERS-CoV in camels, Kenya, 2016–2017. Preventive Veterinary Medicine, 185, 105197. 10.1016/j.prevetmed.2020.105197

